# Evidence of epistasis in regions of long-range linkage disequilibrium across five complex diseases in the UK Biobank and eMERGE datasets

**DOI:** 10.1101/2022.10.19.22280888

**Authors:** Pankhuri Singhal, Yogasudha Veturi, Scott M. Dudek, Anastasia Lucas, Alex Frase, Steven J. Schrodi, David Fasel, Chunhua Weng, Rion Pendergrass, Daniel J. Schaid, Iftikhar J. Kullo, Ozan Dikilitas, Patrick M.A. Sleiman, Hakon Hakonarson, Jason H. Moore, Scott M. Williams, Marylyn D. Ritchie, Shefali S. Verma

**Affiliations:** Department of Genetics, Perelman School of Medicine, University of Pennsylvania, Philadelphia, PA 19104; Laboratory of Genetics, School of Medicine and Public Health, University of Wisconsin, Madison, WI 53706; Columbia University, New York, NY 10027; Genentech, San Francisco, CA 94080; Mayo Clinic, Rochester, MN 55902; Children’s Hospital of Pennsylvania, Philadelphia, PA 19104; Department of Computational Biomedicine, Cedars-Sinai Medical Center, Los Angeles, CA 90048; Department of Genetics and Genome Sciences, School of Medicine, Case Western Reserve University, Cleveland, OH 44106; Department of Pathology and Laboratory Medicine, Perelman School of Medicine, University of Pennsylvania, Philadelphia, PA 19104

**Keywords:** epistasis, linkage disequilibrium, long-range, complex human disease, variable expressivity, pleiotropy, essential genes, evolution, interchromosomal

## Abstract

Leveraging linkage disequilibrium (LD) patterns as representative of population substructure enables the discovery of additive association signals in genome-wide association studies (GWAS). Standard GWAS are well-powered to interrogate additive models; however, new approaches are required to investigate other modes of inheritance such as dominance and epistasis. Epistasis, or non-additive interaction between genes, exists across the genome but often goes undetected due to lack of statistical power. Furthermore, the adoption of LD pruning as customary in standard GWAS excludes detection of sites in LD that may underlie the genetic architecture of complex traits. We hypothesize that uncovering long-range interactions between loci with strong LD due to epistatic selection can elucidate genetic mechanisms underlying common diseases. To investigate this hypothesis, we tested for associations between 23 common diseases and 5,625,845 epistatic SNP-SNP pairs (determined by Ohta’s *D* statistics) in long-range LD (> 0.25cM). We identified five significant associations across five disease phenotypes that replicated in two large genotype-phenotype datasets (UK Biobank and eMERGE). The genes that were most likely involved in the replicated associations were 1) members of highly conserved gene families with complex roles in multiple pathways, 2) essential genes, and/or 3) associated in the literature with complex traits that display variable expressivity. These results support the highly pleiotropic and conserved nature of variants in long-range under epistatic selection. Our work supports the hypothesis that epistatic interactions regulate diverse clinical mechanisms and may especially be driving factors in conditions with a wide range of phenotypic outcomes.

**Significance:** Current knowledge of genotype-phenotype relationships is largely contingent on traditional univariate approaches to genomic analysis. Yet substantial evidence supports non-additive modes of inheritance and regulation, such as epistasis, as being abundant across the genome. In this genome-wide study, we probe the biomolecular mechanisms underlying complex human diseases by testing the association of pairwise genetic interactions with disease occurrence in large-scale biobank data. Specifically, we tested intrachromosomal and interchrosomal long-range interactions between regions of the genome in high linkage disequilibrium, these regions are typically excluded from genomic analyses. The results from this study suggest that essential gene, members of highly conserved gene families, and phenotypes with variable expressivity, are particularly enriched with epistatic and pleiotropic activity.

## Introduction

Genome-wide scans serve as a foundation for understanding complex traits by elucidating how genomic variation affects phenotypic variation. However, the nature of biological systems suggests that relationships between genotypes and phenotypes are often more complex than can be detected using the methods usually employed (Moore, 2003). Extant phenotypic variation is a consequence of evolutionary processes and environmental effects, resulting in allele frequency changes within a population (Draghi, 2019). Phenotypic variation explained is due to a combination of additive and non-additive effects that together define broad-sense heritability (Sella & Barton, 2019). Non-additive effects, including higher order interactions or epistasis, can be interpreted as dependencies or complex relationships between genes or other sources of genetic variation that influence the presentation of a phenotype. Studies in model organisms demonstrate that epistatic interactions are a key factor driving phenotypic complexity, but the role of epistasis in phenotypic determination in humans remains elusive (Lehner, 2007; Mackay & Moore, 2014a). When studies that statistically test for interaction of genetic variants do identify higher order interactions, they are often hard to replicate due to reasons such as but not limited to: model instability, insufficient model complexity due to missing variables, limited statistical power in replication datasets, changes in allele frequency, variation in contextual factors, and lack of interpretability of identifiable models (Greene et al., 2009). Hence, the role of non-additive effects in the context of disease mechanisms remains a challenge to elucidate.

Evolution shapes our genomes via natural processes. Changes in genomic structure that can affect disease risk and population disparity are driven by a variety of evolutionary mechanisms such as genetic drift, gene flow, single locus, and polygenic selection (Charlesworth & Charlesworth, 2010). Phenotypes of clinical relevance are often determined by combinations of loci across the genome. Therefore, to dissect the genetic architecture of clinical phenotypes, it is of particular importance to construct models that define relationships imitating evolutionary processes. As model instability and non-interpretability have hindered replication of epistatic interaction for human traits, using evolutionary processes to model genetics of complex traits can enrich our ability to detect epistasis (Mackay & Moore, 2014b).

Evolutionary processes can produce genomic patterns of variation such as linkage disequilibrium (LD), or non-random associations of alleles (Slatkin, 2008). Since the patterns observed are due to past events, leveraging LD patterns to recapitulate evolutionary processes can enhance understanding of biological mechanisms. For example, strong LD (R^2^ >0.8) observed in conserved genomic regions across ancestry groups might indicate a functional relationship among variants that are of fundamental cellular phenomena (Guryev et al., 2006). (Guryev et al., 2006). Loci can remain in strong LD for many reasons, including physical proximity and functionality as a “supergene.” The mode of inheritance known as supergene occurs due to genomic rearrangement that strives to preserve or lock beneficial alleles across more than one gene (Thompson & Jiggins, 2014). This phenomenon of multiple tightly linked loci regulating a system of discrete phenotypes has been observed across the animal kingdom in functionally related genes that clearly contribute to a shared phenotype (Jeong et al., 2022; Joron et al., 2011). Evidence of physical interactions between regions that harbor regulatory elements alludes to the importance of non-additive effects. For example, Miele and Dekker document well-characterized cases of long-range interactions involved in activation and repression of transcription (Miele & Dekker, 2008).

Ohta’s *D* statistics were developed to parse LD in order to determine the contribution of epistasis from population subdivision (Ohta, 1982). By partitioning the LD between a pair of loci into components within and between populations, the components attributable to differences in allele frequencies among subpopulations and to epistatic selection can be estimated. SNPs in strong LD are typically pruned out of genomic analyses to reduce the burden of “redundant” variants (Calus & Vandenplas, 2018). Genome-wide association studies (GWAS) leverage LD by testing associations between phenotypes and tag SNPs, which function as identifiable proxies for causal SNPs. Experiments in model systems have shown that variants under high selection in evolutionarily conserved regions undergo epistatic selection, possibly as a means of genomic regulation (Mackay, 2014). Although many non-coding regions have levels of evolutionary conservation comparable to those of protein-coding regions, they have a higher abundance of small effect-size variants that can have a significant cumulative impact on phenotypes (Hua & Springer, 2018). Other studies hypothesize roles for epistatic selection in these highly conserved regions such as structure, function, and evolution of proteins through physical interactions, as well as changes in long-range regulatory activity by way of three-dimensional chromatin conformation (Huang et al., 2012; Navarro-Dominguez et al., 2022; Schaeffer & Miller, 1993).

In humans, the biological mechanisms related to regions under epistatic selection remain unknown, thus there has been a long-standing debate about the role of epistasis in disease etiology. If genetic systems are assumed to be in flux, then evolution can be thought of as continuously “tinkering” or adding variation to functionally solve problems (Jacob, 1977). Phillips et al. describe this as something of a house of cards, in which the removal of one central component can bring the entire house down, given that one locus may be interacting with many other genes (Phillips, 2008). This intrinsic structural dependency predicated on an iterative accumulation of small changes can be thought of as the byproduct of 3.5 billion years of descent with modification, as opposed to an intricate piecemeal system (Lynch, 2007). Some alleles have stronger effects on the overall genetic system that can be captured as “main effects” in statistical models, whereas other alleles contribute more subtle effects through modes of regulation such as transcription, splicing, and epistasis (Phillips, 2008). Univariate and multivariate approaches with interaction effects can be used in tandem to yield greater insights into genetic disease mechanisms than can be obtained using either approach alone.

There has been little study of the effects of epistatic interactions between regions of long-range LD, especially in cases where the LD spans across chromosomes. Previous work has found an association between tightly linked SNPs in interchromosomal interactions and aging-related phenotypes such as premature death (Kulminski et al., 2013). A comprehensive review by Maass et al. provides background on nuclear architecture and the roles of chromosomal interactions in genome organization and cellular processes (Maass et al., 2019). Genomic interactions in three-dimensional space are influenced by chromosome topology and transcriptional programs (Gandhi et al., 2012; Krueger et al., 2012). Techniques such as CRISPR live-cell imaging and Hi-C have contributed to our understanding of interchromosomal and intrachromosomal interactions (Belton et al., 2012; Clow et al., 2022). Recent work in mouse and human cell models has shown that certain three-dimensional interchromosomal interactions are a prerequisite for proper physiological gene expression programs and therefore exhibit conservation (Maass et al., 2019); Barutcu et al, 2018). For example, the long non-coding RNA locus *CISTR-ACT* on chromosome 12 regulates chondrogenic gene expression via *cis-*acting interactions with *PTHLH* on chromosome 12 and *trans-*acting interactions with the transcription factor *SOX9* on chromosome 17 (Maass et al., 2012a). Non-homologous chromosomal contacts (NHCCs) occur over longer distances and are more likely to be missed in pairwise interaction analyses than homologous chromosomal contacts unless there is a specific focus on long-range interactions. In the case of *CISTR-ACT* and *SOX9*, physical disruption of tissue-specific NHCCs causes local reorganization of the genome, rendering transcriptional programs dysfunctional and leading to a congenital cartilage malformation known as chondrodysplasia brachydactyly type E (Maass et al., 2012b).

The main objective of this study was to uncover associations of disease phenotypes with long-range epistatic interactions between evolutionarily conserved regions in strong LD. Based on the pervasiveness of epistasis and what is known about epistatic selection and interchromosomal interactions, we hypothesize that uncovering long-range, high-LD interactions due to epistatic selection can help uncover genetic mechanisms underlying common diseases (Koch et al., 2013).

## Results

### Study overview

A workflow schematic of the study is provided in **Fig. 1**. From a cohort of 384,331 individuals in the UK Biobank (UKBB), 23 different case/control sets were created for a range of complex diseases based on the PheCode system (Sudlow et al., 2015; W. Q. Wei et al., 2017). Phenotypes from diverse disease domains including cardiovascular, neurological, immune, rheumatic, pulmonary, ocular, gastrointestinal, dermatologic, and neoplastic diseases were selected to investigate the role of epistasis in independent pathologies affecting different tissues. A consideration of minimum case count was also made when selecting phenotypes. The mean age of the individuals in the UKBB dataset was 57.07 years (standard deviation = 8.07; 55.3% female). All of the phenotypes selected for our analysis, except for breast cancer, included both males and females. We tested for associations between the 23 disease phenotypes and 5,625,845 epistatic SNP-SNP models (determined by Ohta’s *D* statistics, described in detail in the *Methods*) that were in long-range LD (> 0.25cM) and conserved across ancestral populations. Associations that were significant after Bonferroni correction were tested for replication in the eMERGE consortium dataset (n = 50,646; 50.5% female; mean age 68.15 years, standard deviation = 18.96 years) (McCarty et al., 2011). All individuals in the eMERGE and UKBB cohorts were of European ancestry. Case/control count information for each phenotype can be found in **Table S1**. Further details of the analysis are provided in the *Methods* section.

**Fig. 1:**
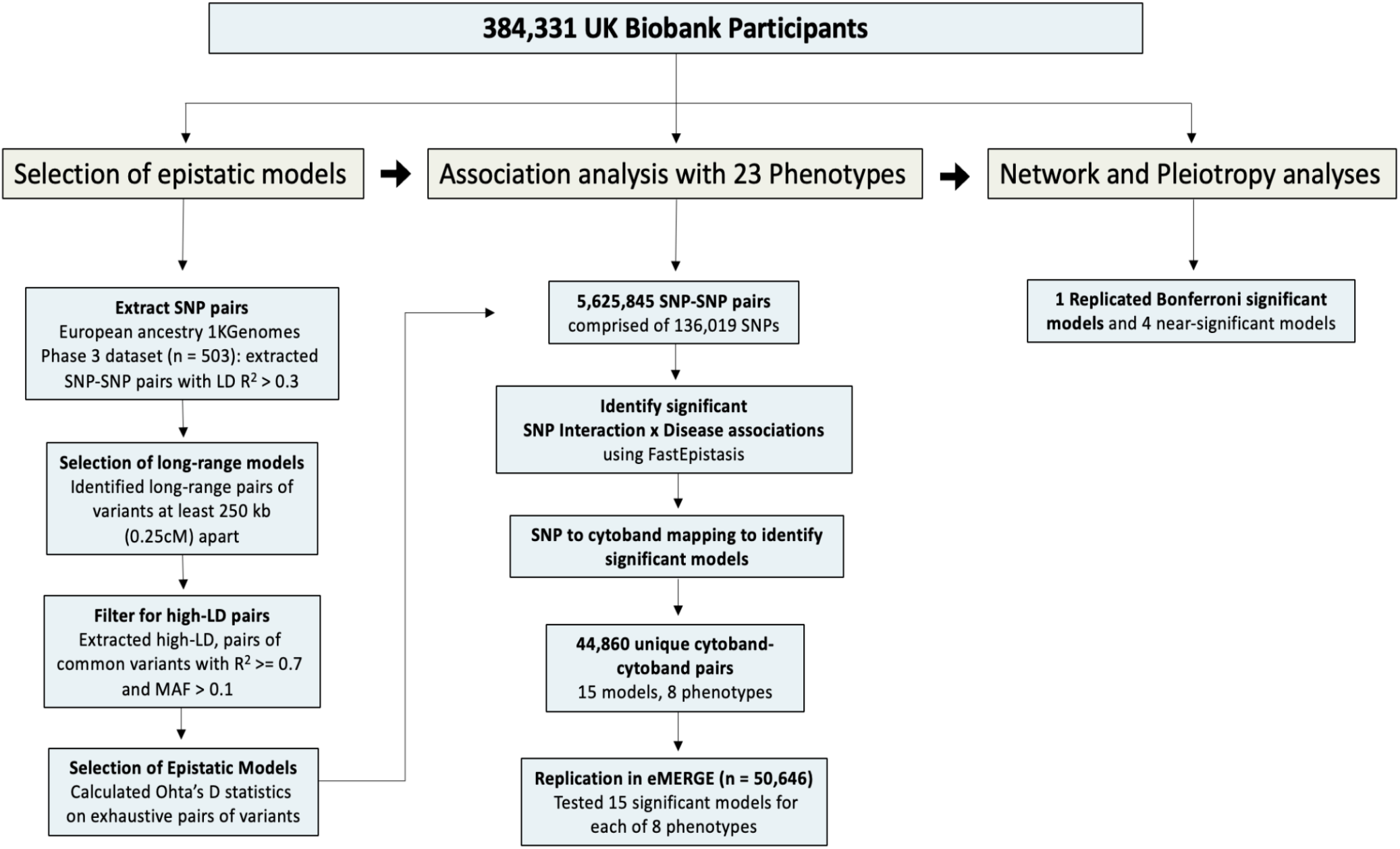
Study workflow. The first part of this study extracted epistatic SNP-SNP interaction models from 1000 Genomes dataset based on Ohta’s *D* statistics. The second part of the study tested associations of each pair of SNPs across 23 complex phenotypes in the UKBB using FastEpistasis. Significant models were tested for replication in the eMERGE dataset. Replicating models were characterized further in the third part of the study.

### Association testing of epistatic interactions with complex phenotypes

A total of 5,625,845 epistatic, long-range, high-LD SNP-SNP models were tested for association with each of 23 different phenotypes in the UKBB dataset using the FastEpistasis test, which models additive by additive SNPs (Schüpbach et al., 2010). We mapped each SNP to a chromosome cytoband region forming what we refer to as a *cytoband-cytoband* (cyto-cyto) model. Each chromosome arm is divided into regions, or cytogenetic bands (cytobands), that can be seen under a microscope when using specific stains (Dolan, 2011). The SNPs mapped to cytobands are labeled according to their distance from the centromere on the p or q arm of the chromosome. We evaluated the epistatic associations under the framework of cyto-cyto models because the underlying hypothesis of epistasis was driven by identifying regions of chromosomal interaction rather than specific participating SNPs. Interpretation of specific SNP-SNP interactions is difficult because of local LD substructures; however, these are implicitly considered in SNP-to-cytoband mapping. After applying Bonferroni correction to adjust for the number of unique cyto-cyto models (0.05 / 44,860 = 1.1 × 10^−6^), the top SNP-SNP model significant for each unique cytoband-cytoband mapping was selected in cases where multiple models reached statistical significance. In the UKBB data, 15 out of 44,860 unique cyto-cyto models spanning 9 of the 23 phenotypes reached statistical significance (**Table 1**). All UKBB FastEpistasis summary statistics can be found in supplemental files.

**Table 1:** SNP-SNP interactions and disease associations identified by FastEpistasis test in the UKBB dataset. Associations between 5,625,845 SNP pairs and 23 phenotypes were tested using FastEpistasis in the UKBB dataset. Bonferroni correction was performed by mapping the SNPs to cytoband regions and adjusting for the total number of unique cytoband-cytoband pairs (0.05 / 44,860 = 1.1 × 10^−6^). Asterisk (*) denotes significant p-values after Bonferroni correction. Shown here are the top SNP-SNP models of each unique chromosomal pairing across all phenotypes.

| Phenotype | CHR A | SNP A | CHR B | SNP B | P-value |
| --- | --- | --- | --- | --- | --- |
| Acute Pulmonary Heart Disease | 12 | 12:6287428 | 10 | 10:93068004 | 9.60E-06 |
| AD | 6 | 6:32648500 | 6 | 6:32364667 | 6.47E-07* |
| COPD | 15 | 15:81121350 | 3 | 3:16237372 | 2.06E-06 |
| T2D | 16 | 16:7788521 | 14 | 14:86160697 | 2.29E-07* |
| Essential Hypertension | 12 | 12:112230036 | 12 | 12:111962581 | 4.89E-06 |
| Fibromyalgia | 20 | 20:45562611 | 13 | 13:39174411 | 1.79E-06 |
| Glaucoma | 9 | 9:133944198 | 7 | 7:119526478 | 7.68E-07* |
| Hepatic Infection | 13 | 13:39174411 | 8 | 8:30793179 | 1.57E-06 |
| Herpes | 1 | 1:78446761 | 1 | 1:78092479 | 2.42E-07* |
| Hyperplasia of Prostate | 15 | 15:38271345 | 4 | 4:24167431 | 1.10E-07* |
| Hyperplasia of Prostate | 17 | 17:66168247 | 8 | 8:63904818 | 2.46E-07* |
| Idiopathic Proctocolitis | 3 | 3:51689306 | 3 | 3:51215148 | 2.14E-06 |
| Iron Deficiency | 1 | 1:201268216 | 1 | 1:201002649 | 4.94E-07* |
| Iron Deficiency | 22 | 22:45594814 | 4 | 4:156510640 | 5.27E-08* |
| Ischemic Heart Disease | 12 | 12:6287428 | 10 | 10:93068004 | 1.06E-05 |
| MDD | 15 | 15:58475374 | 2 | 2:26628863 | 5.68E-06 |
| Cancer of Digestive Organs | 4 | 4:48783587 | 4 | 4:48493237 | 3.48E-06 |
| MS | 19 | 19:49194880 | 19 | 19:13514610 | 2.96E-07* |
| MS | 6 | 6:32648500 | 6 | 6:32364667 | 4.96E-10* |
| Pancreatitis | 20 | 20:45562611 | 13 | 13:39174411 | 1.79E-06 |
| Psoriasis | 12 | 12:114954298 | 9 | 9:130002630 | 4.53E-07* |
| Psoriasis | 14 | 14:86260029 | 5 | 5:58499153 | 2.87E-07* |
| Psoriasis | 4 | 4:79179009 | 3 | 3:134787497 | 1.64E-07* |
| SCZ | 11 | 11:39275165 | 8 | 8:116432183 | 5.53E-07* |
| SCZ | 9 | 9:95674613 | 5 | 5:146534330 | 3.30E-07* |
| Tonsillitis | 3 | 3:164060391 | 3 | 3:163803419 | 1.42E-06 |
| Ulcerative Colitis | 3 | 3:51689306 | 3 | 3:51215148 | 2.17E-06 |
| Viral Infection | 12 | 12:105965951 | 6 | 6:67707841 | 2.76E-06 |
| Breast Cancer | 4 | 4:155839500 | 4 | 4:150941758 | 1.02E-05 |

We next tested all SNP-SNP pairs mapping to each of the 15 significant cyto-cyto models for associations with their corresponding phenotypes in the eMERGE dataset. The Bonferroni corrected threshold for eMERGE was adjusted for the 15 cytoband-cytoband models tested (p-value = 0.05 / 15 = 3.3 × 10^−3^). Only one of the 15 cyto-cyto models reached statistical significance in the eMERGE data. This model was associated with type 2 diabetes (T2D; chromosome 16:p13.3 × chromosome 14:q31.3; p-value = 0.001344; **Table 2**). Several other models came close to statistical significance: two associated with multiple sclerosis (MS; chromosome 19:q13.33 × chromosome 19:p13.2 and chromosome 6:p21.32 × chromosome 6:p21.32; p-values = 0.003466 and 0.003616, respectively), one associated with psoriasis (chromosome 14:q31.3 × chromosome 5:q11.2; p-value = 0.004205), and one associated with schizophrenia (SCZ; chromosome 11:p12 × chromosome 8:q23.3; p-value = 0.007327). The significant and nearly significant models are referred to hereafter as “top models” and are shown in **Table 2**. A graphical representation of the top models is shown in **Fig. 2**, highlighting the intrachromosomal and interchromosomal interactions between genes in the cytoband regions. All eMERGE top model FastEpistasis summary statistics can be found in supplemental files.

**Table 2:** Cytoband-cytoband models that were significant or nearly significant in both the UKBB and eMERGE datasets. The SNP-SNP model with the lowest p-value from each unique cytoband-cytoband bin was tested in the eMERGE dataset if it met Bonferroni significance in the UKBB dataset. A total of 15 models (* in **Table 1**) were tested for replication, of which one reached significance in the eMERGE dataset (**) and four reached near-significance (*) at a significance threshold of P = 3.3 × 10^−3^.

|  |  |  | UKBB |  |  |  |  | eMERGE |  |  |  |  |
| --- | --- | --- | --- | --- | --- | --- | --- | --- | --- | --- | --- | --- |
| Phenotype | Cyto A | Cyto B | SNP A | Gene A | SNP B | Gene B | P-value | SNP A | Gene A | SNP B | Gene B | P-value |
| T2D | p13.3 | q31.3 | 16:7788521 | RBFOX1 | 14:86160697 | FLRT2 | 2.29E-07 | 16:6426717 | RBFOX1 | 14:86186113 | FLRT2 | 0.001344** |
| MS | q13.33 | p13.2 | 19:49194880 | FUT2 | 19:13514610 | CACNA1A | 2.96E-07 | 19:49196722 | FUT2 | 19:13514610 | CACNA1A | 0.003466* |
| MS | p21.32 | p21.32 | 6:32648500 | HLA-DQB1 | 6:32364667 | BTNL2, HCG23 | 4.96E-10 | 6:32665629 | MTCO3P1 | 6:32397863 | BTNL2 | 0.003616* |
| Psoriasis | q31.3 | q11.2 | 14:86260029 | FLRT2 | 5:58499153 | PDE4D | 2.87E-07 | 14:85403658 | LOC100421611 | 5:58499153 | PDE4D | 0.004205* |
| SCZ | p12 | q23.3 | 11:39275165 | RPL18P8 | 8:116432183 | TRPS1 | 5.53E-07 | 11:41251442 | LRRC4C | 8:116351905 | TRPS1 | 0.007327* |

**Fig. 2:**
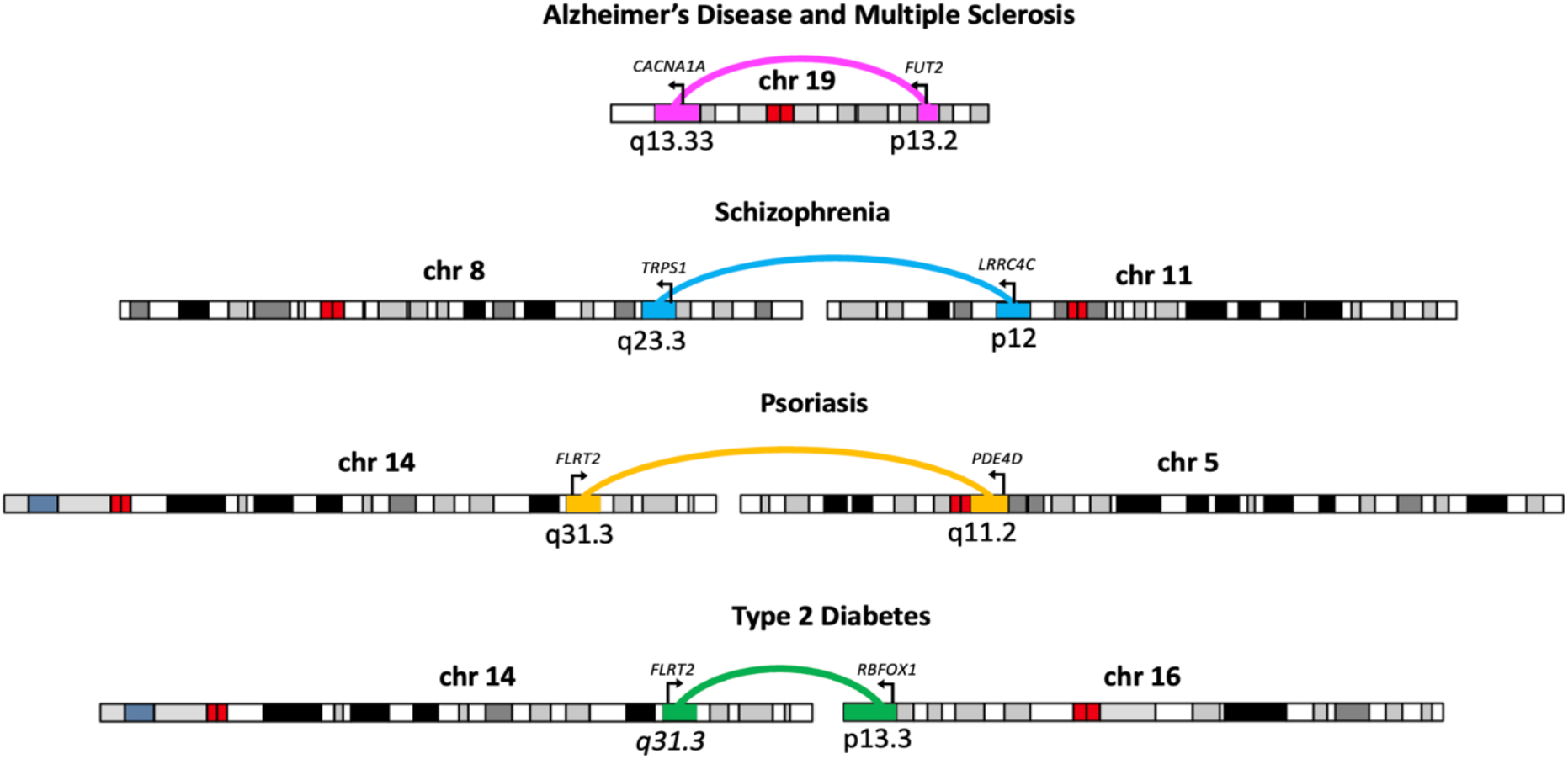
Models that were significant based on UKBB data and were replicated in the eMERGE data. Interchromosomal and intrachromosomal interactions and disease associations of the top models that replicated in both datasets are shown.

The MS intrachromosomal model for chromosome 6:p21.32 mapped to the *HLA* region (chromosome 6:p21.3), which is well characterized in the literature for its dynamic role encoding cell-surface proteins responsible for regulation of the immune system (Dendrou et al., 2018). MS, as well as numerous other conditions including Alzheimer’s disease (AD), type 1 diabetes, and rheumatic heart disease, have been linked to the *HLA* region (Auckland et al., 2020; Jiang et al., n.d.; R.-C. Lu et al., 2017; Patsopoulos et al., 2013). This highlights the strong genetic effect that combinations of alleles in the *HLA* region have on disease susceptibility and protection. These results are promising and may serve to substantiate previous findings; however, a focus on results on the *HLA* region is out of the scope of this study, given the current challenges in sequencing the *HLA* region for interpretation.

### Univariate analysis to test if interacting SNPs function as main effects

Next, individual SNPs in each significant cyto-cyto model were tested in a univariate regression framework to determine if they function as main effects for their respective phenotype(s). In addition to the SNPs used to determine the p-values for the cyto-cyto models, we tested any proxy SNPs that were also in LD (R^2^ < 0.5) and within 1MB upstream or downstream of the representative SNPs. The plots in **Fig. S1 A–J** depict the results of the univariate analyses of the SNPs in each model, with the chromosome with lowest p-values for each model shown. Bonferroni correction was based on the number of proxy SNPs tested. No statistically significant main effects were found, although the results for chromosome 6 overlapped with GWAS hits for various traits, as expected given the dynamic nature of the HLA region. Summary statistics for main effects analysis can be found in supplementary files.

### Network analysis to predict molecular mechanisms linking epistatic gene pairs in MS

To probe molecular mechanisms that might explain the epistatic interactions between the top gene pairs from our analysis, we created biological process gene networks using HumanBase (Greene et al., 2015). We selected the biological process terms that best described each gene in a pair one at a time to generate molecular networks of interaction. For each gene in the network, a score was generated to indicate the average weight of connections to the epistatic gene pair, subsequently referred to as query genes. The networks in which the query genes had the highest connection scores were evaluated further, with focus given to first- and second-degree neighbors interacting with the query genes. **Fig. 3** shows three biological process gene networks for the intrachromosomal interaction between *FUT2* and *CACNA1A* (chromosome 19). *CACNA1A* is part of a family of genes that provide instructions for making calcium channels, so we generated networks for both “calcium ion transport” and “calcium-mediated signaling” pathways. *FUT2* and *CACNA1A* both had connection scores of 0.46 in the calcium ion transport network and 0.24 in the calcium-mediated signaling network, suggesting that calcium ion transport better describes the functional context of their interaction. This is further reflected by the high number of high-confidence interactions between primary neighbor nodes connected to both genes in the calcium ion transport network, in contrast to the lower number and lower confidence of interactions between primary neighbor nodes and query genes in the calcium-mediated signaling network. Because *FUT2* is responsible for the composition and functional properties of glycans in bodily secretions, we also generated a “proteoglycan biosynthetic process” network; however, the low confidence scores and the low number of primary neighbor nodes connecting both genes in this network do not seem to support a molecular interaction hypothesis for *FUT2* and *CACNA1A* in the proteoglycan biosynthetic process as strongly.

**Fig. 3:**
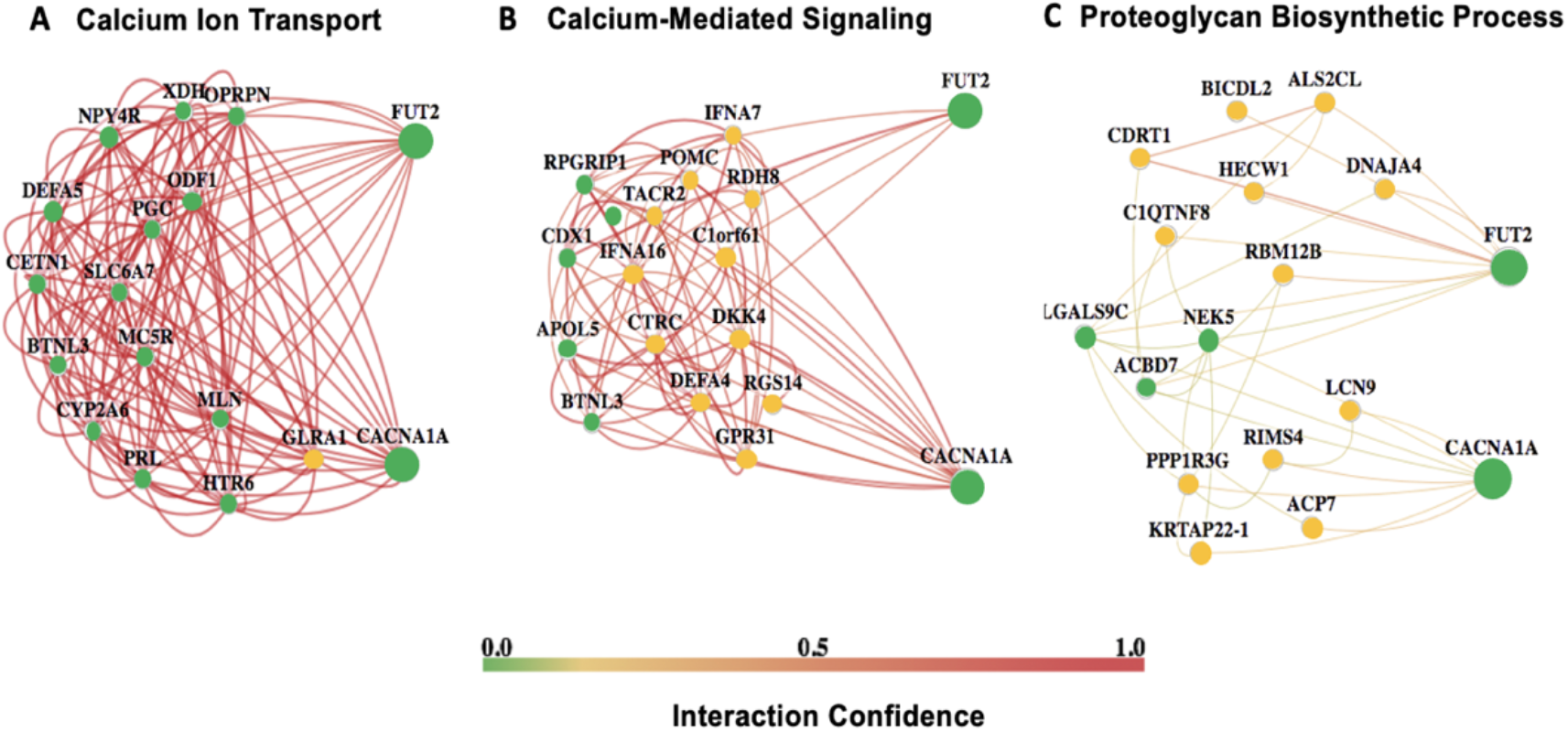
A biological function network analysis of *FUT2* and *CACNA1A* interaction. Biological function networks are shown for different selected processes potentially involving the *FUT2* × *CACNA1A* interaction (associated with MS and AD). The interaction confidence scale reflects the strength of the edge weights. *FUT2* and *CACNA1A* are both green nodes, as well as all nodes directly connected to both query genes (primary neighbors). Yellow nodes are secondary neighbors, connected to one query gene or the other. **A**. The calcium ion transport process has many first-degree nodes common to *FUT2* and *CACNA1A*, as well as high-confidence edges, suggesting a strong fit as a process common to both genes. **B**. The calcium-mediated signaling network has more secondary neighbors, suggesting a less strong fit. **C**. The proteoglycan biosynthetic process network has few primary neighbors to both genes, and the edges have low interaction confidence, suggesting this is less likely to be a biological process linking *FUT2* and *CACNA1A*.

## Discussion

We hypothesized that long-range epistatic interactions in chromosomal regions with high LD are implicated in mechanisms underlying complex diseases. To test this, we looked for associations between 23 complex disease phenotypes and 5,625,845 epistatic pairs of SNPs with strong, long-range LD. Although we tested associations for specific pairs of SNPs, we interpreted the results in terms of the cytoband regions containing the SNPs. There is substantial debate in the genomics community about how to best link non-coding and intergenic SNPs to corresponding functional genes. Our approach using cytoband regions enabled us to make functional hypotheses about non-coding or intergenic SNPs based on the nearby genes that were most likely to have significant effects on the phenotypes in question.

One interesting finding was that none of the SNPs in our top models had significant effects on the phenotypes of interest in univariate analyses (**Fig. S1**), indicating that the phenotypic associations were driven by interactions between the SNPs rather than by the main effects of each SNP alone. The genes linked to top models seem to be particularly enriched for dynamic roles as part of large, conserved gene families active during development.

### Chromosomes 16p13.3 and 14q31.3 in type 2 diabetes

Our FastEpistasis analysis identifies the top replicating model to be an interchromosomal interaction between *FLRT2* (chromosome 14q31.3) and *RBFOX1* (chromosome 16p13.3), associated with T2D. The interacting SNPs in the UKBB dataset map to an intergenic region 66,427 bp downstream of *FLRT2* and an intergenic region 25,181 bp downstream of *RBFOX1*, respectively. *RBFOX1*, one of three mammalian paralogs of the *RBFOX* gene family, regulates tissue-specific alternative splicing and post-transcriptional regulation in the brain and heart (Damianov et al., 2016). *FLRT2* is part of the FLRT gene family encoding membrane proteins involved in the regulation of cell adhesion and repulsion, cell migration, cell signaling, and axon guidance (Li et al., 2021a). *RBFOX1* has been associated with neurodegenerative and cardiometabolic traits (Zhao, 2013). Previous studies support a role for *RBFOX1* in regulating beta cell gene expression through neuron-like alternative splicing regulation (Juan-Mateu et al., 2017a; Nutter et al., 2017). One explanation for this could be because the pancreas is highly innervated, causing increased neuron-specific transcriptional programs. Another could be that neurons share many phenotypic traits with pancreatic beta cells since both use similar exocytotic machinery to secrete insulin and neurotransmitters (Juan-Mateu et al., 2017;Arntfield & van der Kooy, 2011). Given that FLRT2 is known to interact with proteins such as ADGRL3, FGFR2, and UNC5D to mediate various cell signaling pathways, we hypothesize that the interaction between chromosomes 14 and 16 regulates transcriptomic activity in T2D through an alternative splicing program (K. Wei et al., 2011).

### Chromosome 5q11.2 and chromosome 14q31.3 in psoriasis

*FLRT2* was also found to affect psoriasis through an interaction with the intronic region of *PDE4D* on chromosome 5q11.2. PDE4D is part of the cyclic nucleotide phosphodiesterase family of enzymes that hydrolyze the intracellular second messenger cAMP, a key signal transduction molecule in numerous biological processes (Ong et al., 2009). Patients with psoriasis display overexpression of *PDE4D* mRNA in peripheral blood mononuclear cells compared with control individuals (Schafer et al., 2016). Apremilast, an oral small-molecule inhibitor of PDE4D, is used to treat psoriasis and other chronic inflammatory disorders such as asthma and Behçet’s disease through inhibition of the Th17 pathway (Afra et al., 2019; Chen et al., 2020; Schett et al., 2010). Our results corroborate the established proinflammatory link between *PDE4D* and psoriasis; however, the mechanistic basis for a *PDE4D* and *FLRT2* interaction association with psoriasis is unknown.

*FLRT2* functions as a cell-surface signaling protein that interacts dynamically with various proteins during developmental events, especially axon guidance (Li et al., 2021b). Given the role of *FLRT2* in modulating cortical migration during nervous system development, Akita et al. proposed an analogous guidance function for *FLRT2* in vascular development (Akita et al., 2016). *FLRT2* is one of the primary ligands that binds and activates the adhesion G protein-coupled receptor *LPHN2*, which acts as a repulsive guidance receptor that controls blood vessel structure and function in model systems (Camillo et al., 2021). *FLRT2* has also been identified as an autoantigen, or cell-surface target, of anti-endothelial cell antibodies in the vascular systems of patients with systemic lupus erythematosus (Shirai et al., 2012). Once *LPHN2* is activated by *FLRT2* it elicits the synthesis of cAMP, which is hydrolyzed by PDE4D (Ong et al., 2009; Sando & Südhof, 2021). Our results together with the previous findings suggest that interaction between *PDE4D* and *FLRT2* has a proinflammatory effect in patients with psoriasis, potentially acting through an autoimmune pathology in the vascular system. Further functional experiments are needed to understand the mechanism.

### Chromosome 8q23.3 and chromosome 11p12 in schizophrenia

We found that an interaction between *TRPS1* (chromosome 8q23.3) and *LRRC4C* (chromosome 11p12) is associated with schizophrenia. The SNP on chromosome 8 is 531,181 bp upstream of *TRPS1*, whereas the SNP on chromosome 11 falls in intron 1 of the *LRRC4C* gene. TRPS1 is a transcriptional repressor that binds to GATA-regulated genes during different stages of embryonic development to influence chondrocyte proliferation and differentiation (Wang et al., 2018). *LRRC4C* encodes a post-synaptic adhesion molecule that binds with the conserved family of netrin G ligand (NGL) proteins (Choi et al., 2019) to regulate synaptic organization. There is no previous evidence that *TRPS1* and *LRRC4C* interact to influence schizophrenia; however, *LRRC4C* has been implicated in brain disorders including schizophrenia, bipolar disorder, autism spectrum disorder, and developmental delay (Maussion et al., 2017; Zhang et al., 2021). Mouse models have shown that mice lacking NGL-1 exhibit hyperactivity and anxiolytic-like behavior due to widespread excitation of neurons in the brain. This suggests that *LRRC4C* plays a role in suppression or dampening of neuronal activity (Choi et al., 2019). We hypothesize that *TRPS1* acts as a repressor of *LRRC4C*, which is supported by a previous analysis of GTEx bulk tissue expression data (GTEx Consortium, 2013) showing that *LRRC4C* (ENSG00000148948.7) is highly expressed in brain tissues, whereas *TRPS1* (ENSG00000104447.12) is expressed at low levels in brain tissue compared with all other tissues.

### Chromosome 19p13.2 and 19q13.33 in multiple sclerosis

An intrachromosomal interaction between *FUT2* (chromosome 19p13.2) and *CACNA1A* (chromosome 19q13.33) was found to be associated with MS. The *FUT2* gene determines blood group secretor status. Being homozygous for the inactive “non-secretor” allele confers susceptibility and resistance to certain infections (Azad et al., 2018). *FUT2* non-secretor status has been shown to result in significantly increased lymphocyte infiltration levels during infection (Santos-Cortez et al., 2018). Previous work highlights the role of lymphocyte-mediated calcium influx patterns observed in autoimmune conditions, including MS (Orbán et al., 2013). Thus, a genetic interaction between *FUT2* and *CACNA1A* could inform a calcium-dependent autoimmune mechanism. We conducted post-hoc network analysis using HumanBase to identify pathways or functions that would explain the context of the molecular interaction between *CACNA1A* and *FUT2* (**Fig. 2**). The gene connections in networks generated for known pathways involving each gene (calcium ion transport, calcium-mediated signaling, and proteoglycan biosynthetic process) suggested that the calcium ion transport network best explains the *FUT2* and *CACNA1A* interaction. The primary neighbors connecting both genes in the calcium ion transport network provide supporting evidence for etiologic hypotheses of MS. *BTNL3*, which has also been associated with AD, was previously found to be associated with high-density lipoprotein (HDL) cholesterol levels (Liu et al., 2022). In another study, high HDL cholesterol levels were associated with reduced brain atrophy and demyelination in MS (Blumenfeld Kan et al., 2019). Unexpectedly, the calcium ion transport network also included *OPRPN* and *ODF1. OPRPN* encodes the PROL1 protein, which functions in penile erection. *ODF1* encodes the protein that forms the outer dense fibers surrounding the sperm tail, which are essential for maintaining the elastic sperm tail structure (Yang et al., 2012). Variation in *OPRPN* and *ODF1* is linked to infertility in men. A previous longitudinal study showed an association between male infertility and MS, which was likely caused in part by a shared genetic component of both conditions (Glazer et al., 2018). These results suggest that interaction between *FUT2* and *CACNA1A* might explain male infertility in MS. Another gene in the calcium ion transport network, *MC5R*, encodes the melanocortin 5 receptor, which exerts immunomodulatory effects by converting primed T cells to regulatory T (Treg) cells (Taylor & Namba, 2001). Reduced Treg cell signaling in chronic inflammation can lead to an increase in the number of autoimmune antigen-presenting cells that ultimately cause a self-destructive central nervous system environment in MS. Our association test does not define a direct link between *MC5R* and MS; however, the network analysis of *FUT2* and *CACNA1A* supports *MC5R* as a potential molecular connection underlying the epistatic effects of *FUT2* and *CACNA1A* on MS.

## Pleiotropy

Pleiotropy, whereby one gene or mutation influences multiple distinct and seemingly unrelated phenotypic traits, has been extensively modeled in animal systems and studied in humans using statistical models. Recent work has found pleiotropy to be a highly prevalent, if not ubiquitous, phenomenon in human genotype-phenotype mapping (Chesmore et al., 2018). Tyler et al. provide a comprehensive review of the relationship between epistasis in pleiotropy, citing that these phenomena are not isolated incidents. Rather, they are inherent properties of biomolecular networks that are critical to the understanding of genetics underlying common human disease and should be modeled extensively (Tyler et al., 2009). Despite substantial evidence, there is no empirical basis for pleiotropy in humans (Paaby & Rockman, 2013; Sivakumaran et al., 2011). Our analysis yields top interaction models that are associated with multiple phenotypes, suggesting a potential pleiotropic etiology for certain clinical pathologies. We further hypothesize molecular mechanisms that could explain our results supporting epistatic and pleiotropic relationships.

### Chromosome 19 in Alzheimer’s disease and multiple sclerosis

Chromosome 19 has long been linked to AD in the GWAS literature by way of variation in genes such as *APOE* (19q13.32), *ABCA7* (19q13,3), and *CD33* (19q13.41) (Moreno-Grau et al., 2018). We found chromosome 19 to play a pleiotropic role in mediating AD and MS through epistatic interactions.

A long-range, intrachromosomal interaction between chromosome 19q13.33 and chromosome 19p13.2 (chr19:13514610) was associated with both AD and MS in our FastEpistasis analysis. In the UKBB dataset, the association of this model with MS (**Table 1)** was statistically significant, whereas the association with AD was nearly significant with p-value = 1.79 × 10^−6^ (**Supplementary files**). In eMERGE, the model associated with MS with a near significant p-value (Table 2) but associated with AD with statistical significance of p-value = 1.27 × 10^−4^ (Supplementary files). The chromosome 19p13.2 variant maps to an intron of *CACNA1A*, which belongs to a highly conserved gene family that provides instructions for making calcium channels. *CACNA1A* is one of the strongest known genetic risk factors for a variety of neurodevelopmental and neurodegenerative conditions including familial AD, trinucleotide repeat conditions, and SCZ; however, its role in the pathogenesis of these conditions is largely unknown (Grosso et al., 2022; Psychiatric GWAS Consortium Bipolar Disorder Working Group, 2011). Located within 2MB upstream of *CACNA1A* are *TRMT1* and *LDLR*, which are both linked to AD and MS (Gopalraj et al., 2005; Y. Lu et al., 2022; Mailleux et al., 2017). The SNP in the intron of *CACNA1A* might regulate that gene in tandem with the SNP on chromosome 19q13.33, or the interaction might be regulating other genes via longer-range effects. The SNP on chromosome 19q13.33 falls in an intergenic region between *FUT2* and *SEC1P. FUT2* has been associated with vitamin B deficiency, cholesterol levels, type 1 diabetes, and dysregulated gut microbiota, all of which have been linked to AD and MS (Blumenfeld Kan et al., n.d.-b; Ellinghaus et al., 2012; Hazra et al., 2008; Najafi et al., 2012; Schepici et al., 2019; Smyth et al., 2011). Previous work has aimed to detect overlap between autoimmune pathways related to MS pathology and blood lipid pathways related to AD pathology, but no common mechanism for MS and AD pathologies has been identified (Podbielska et al., 2021). We hypothesize that a shared cardiometabolic and autoimmune molecular etiology underpins both AD and MS. Functional experiments are needed to elucidate the mechanism.

### Chromosome 14q31.3 in type 2 diabetes and psoriasis

Chromosome 14q31.3 encodes *FLRT2, a* gene known to play a role in cell-cell adhesion, cell migration, and axon guidance. Our findings implicate *FLRT2* in two interchromosomal interactions, a psoriasis-associated interaction with *PDE4D* (chromosome 5) and a T2D-associated interaction with *RBFOX1* (chromosome 16). While the shared etiology of diabetes and psoriasis is unknown, a breadth of work has shown overlap between T2D and psoriasis by way of shared lipid abnormalities, heightened insulin resistance, and cardiovascular risk biomarkers (Brazzelli et al., 2021). To evaluate if this region is predisposed to pleiotropic activity, we determined if other Mendelian traits and syndromes are linked to genes within 14q31.3 based on the Online Mendelian Inheritance in Man (OMIM) database, shown in **Fig. 4** (Hamosh et al., 2005).

**Fig. 4:**
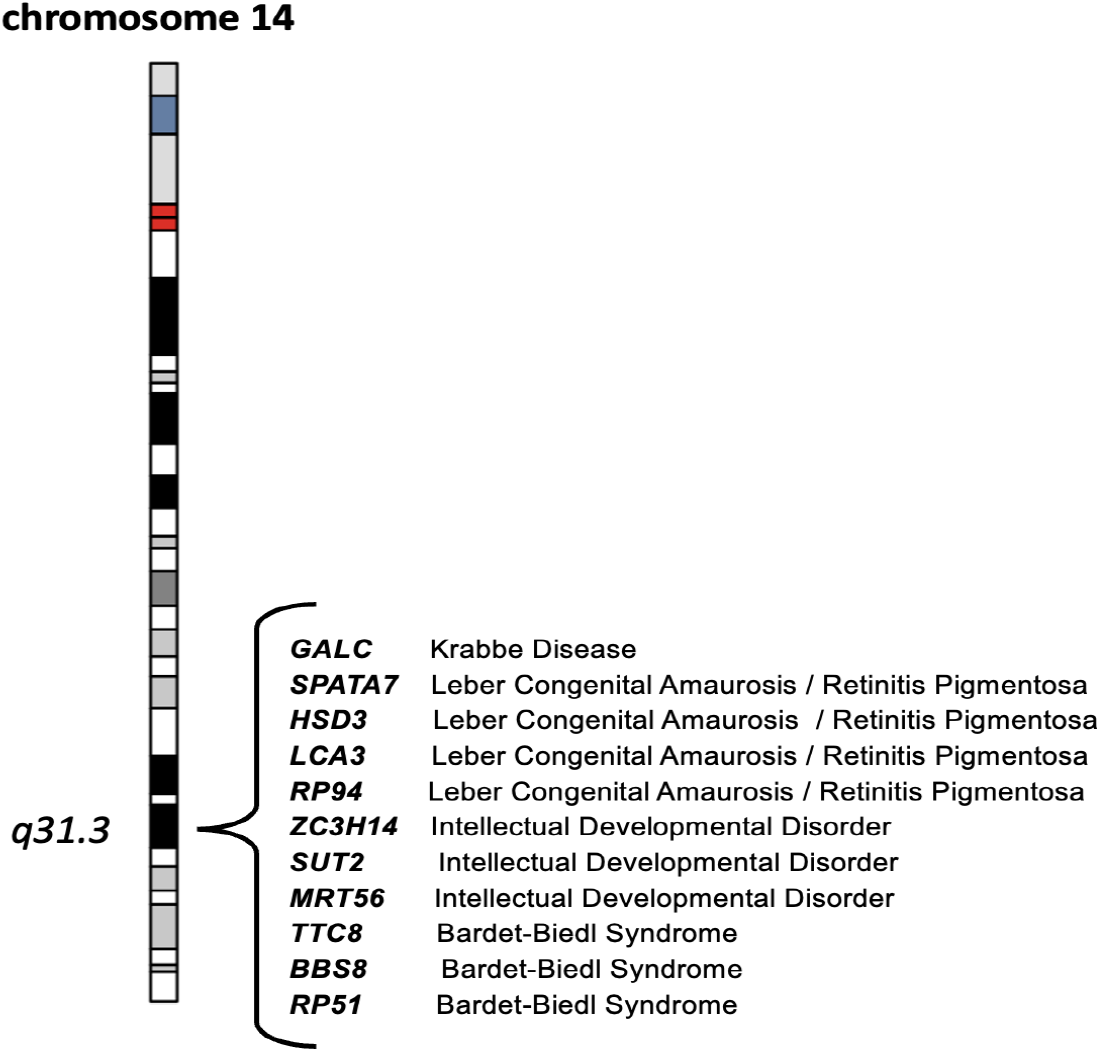
Evidence of pleiotropy on chromosome 14q31.3. Chromosome 14q31.3 interacts with chromosome 16p13.3 to affect T2D and with chromosome 5q11.2 to affect psoriasis. To determine a clinical basis for the pleiotropy observed on chromosome 14q31.3, we used Online Mendelian Inheritance in Man (OMIM) to identify Mendelian conditions known to be associated with this region.

Bardet-Biedl syndrome (BBS) is a pleiotropic ciliopathy with a wide range of clinical variability. While BBS individuals have a high propensity towards T2D due to obesity, it is unclear whether diabetes is a comorbidity or a risk independent of obesity. However, recent mouse models of BBS do support dysregulation of the immune and hematopoietic systems as obesity-independent drivers of T2D (Tsyklauri et al., 2021). Krabbe disease, a rare autosomal recessive lysosomal storage disease, has phenotypic overlap with MS in that its hallmark is demyelination caused by the buildup of unmetabolized lipids throughout the central nervous system (Mar & Noetzel, 2010). Retinitis pigmentosa (RP), a genetic retinopathy that causes vision loss over time, presents varying type of vision loss (e.g., night vision, central vision, or color vision) and severity in different individuals. How lipid dysregulation plays a role in RP disease pathology is not well understood. A previous study found RP to be a result of abetalipoproteinemia (Berson, 2000; Gouras et al., 1971). An understanding of energy metabolism in retinal cells is critical to uncovering the vascular changes that drive the different stages of RP (Fu et al., 2019). Leber congenital amaurosis (LCA) is a group of rare monogenic diseases that frequently result in rapid or progressive vision loss accompanied by other symptoms such as intellectual disability, hearing loss, and cataracts. Lipid changes are not known to be associated with LCA.

The genetic links between chromosome 14q31.3 and BBS, Krabbe disease, RP, and LCA suggest that lipid dysregulation may be the mechanism connecting psoriasis and T2D, as well as playing a role in driving some of these conditions. We hypothesize that epistatic interactions can “modify” and manifest variable expressivity such that the nature of the same condition can vary among individuals. For example, some individuals with BBS who have alleles supporting an interaction involving *FLRT2* may have a higher risk of developing diabetes than individuals with BBS who do not have those alleles. We can only speculate as to why alleles on chromosome 14q31.3 might predispose individuals to cardiometabolic dysregulation in the context of complex and Mendelian conditions. However, our data support the hypothesis that genetic heterogeneity manifests as phenotypic heterogeneity in the context of epistatic interactions.

### Epistasis in essential gene families

To further understand epistatic selection in the context of human disease, we asked whether the genes in our top models function as essential genes, given that essential genes are highly conserved and function in numerous pathways. Previous long-range epistasis work provides a basis for interactions between high-LD SNPs to be thought of as integral to essential biological processes (Kulminski et al., 2013). Essential genes are critical for the survival of organisms under most conditions and commonly drive cell growth and proliferation. The vast majority of human genes are non-essential but still confer some degree of selective advantage. Out of 19,850 known human genes, 3,915 are considered essential genes (Ji et al., 2016).

Essential genes are likely to encode hub proteins that are widely expressed in most tissues, making them well-suited as dynamic disease genes that are active in the interactome (Goh et al., 2007). Based on a previous study of essential genes by Ji et al. and the Full Spectrum of Intolerance to Loss-of-Function (FUSIL) database, six of the seven genes in our top models, *LRRC4C* being the exception, have been classified as essential genes (Cacheiro et al., 2020; Ji et al., 2016). **Fig. 5** depicts the essential genes, categorized by their functional designations from FUSIL, as well as their essential functions in growth and development. Loss of *RBFOX1* or *TRPS1* is developmentally lethal, whereas loss of *FUT2* is non-lethal but results in phenotypic abnormality. When we evaluated the diseases associated with each essential gene, we found that all the essential genes except *FLRT2* are associated with at least two or more related conditions that differ only by a subset of symptoms. A prime example is *CACNA1A*, which is implicated in a variety of neurological conditions ranging from autism spectrum disorder and cerebellar atrophy to epileptic encephalopathy (Damaj et al., 2015). Similarly, *FUT2* has been associated with inflammatory bowel conditions including Crohn’s disease and ulcerative colitis (Wu et al., 2017). Syndromes are sets of complex symptoms that often co-occur, indicating a specific condition. We hypothesize that epistatic variation is a driving factor that differentiates certain sets of overlapping symptoms into distinct pathologies, much like syndromes.

**Fig. 5:**
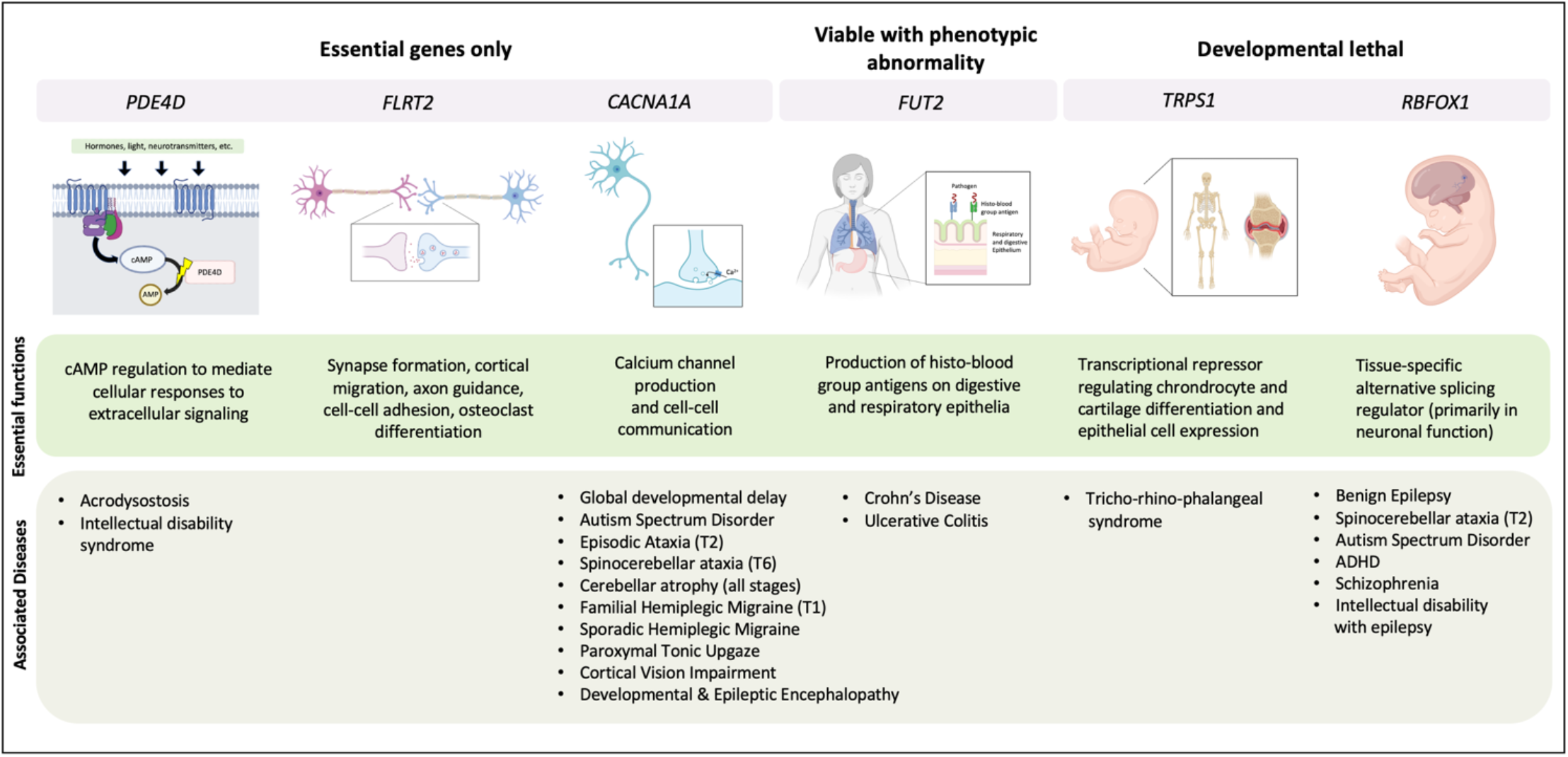
Epistatic interactions map to essential gene families. The biological functions and diseases associated with essential genes in our top models are shown.

Epistatic selection may be occurring at higher rates among essential genes, giving rise to diverse phenotypic outcomes. Given the fundamental cellular roles that essential genes play during and after development, epistasis may be a highly conserved mode of genetic regulation, especially in the context of disease etiology. Functional studies are needed to test the molecular interactions of the essential genes in our top models to further characterize their role beyond development and in adulthood disease.

## Conclusion

We leveraged Ohta’s *D* statistics to 1) identify epistatic interactions between distant regions of the genome that exhibit strong LD, which are typically excluded from genomic analyses, and 2) test association of these interactions with a range of complex diseases. After testing associations of 5,625,845 SNP-SNP pairs with complex diseases, we identified five interactions with disease associations that replicated in the UKBB and eMERGE datasets (**Table 2**). Most of the interactions were interchromosomal, which is surprising given that most long-range genomic and high-throughput chromosome conformation analyses are limited to single chromosomes (Park, 2019). Associations between specific epistatic interactions and conditions including T2D, psoriasis, MS, schizophrenia, and AD were identified. (**Fig. 2**). Furthermore, we identified epistatic interactions with a pleiotropic basis. In particular, chromosome 14:q31.3 had long-range interactions with chromosome 5 (associated with psoriasis) and chromosome 16 (associated with T2D). We conclude that psoriasis and T2D likely share an etiology based on dysregulation of lipid metabolism with an autoimmune component. Our post-hoc analysis of chromosome 14q31.3 showed that four other Mendelian conditions with lipid dysregulation as a potential shared mechanism are also associated with the same cytoband (**Fig. 4**). We conclude from these findings that epistatic interactions may function at a subtle level to modify phenotypic presentation. For example, some individuals with the interaction between chromosome 14 and chromosome 5 may present a lipid-autoimmune phenotype leading to psoriasis. Similarly, individuals with the interaction between chromosome 14 and chromosome 16 may present a lipid-cardio metabolic phenotype leading to T2D, whereas other individuals may present lipid-ocular conditions such as RP. Different variants within the interacting genes may function differently, adding another layer of fine modulation. This is just one example of how epistatic genetic variation might drive variation of disease phenotypes.

Our epistatic gene models appeared to be enriched with members of highly conserved gene families with dynamic and essential roles. For example, *TRPS1* has been associated with a rare autosomal condition called Tricho-rhino-phalangeal syndrome type 1 (**Fig. 5**), which causes highly variable symptoms including hypermobility, craniofacial abnormalities, and hyperhidrosis (Seitz et al., 2001). We hypothesize that epistatic gene interactions modulate the combination of symptoms which are expressed in among individuals. Our results suggest that epistasis can be thought of as ‘fine-tuning’, in which interactions manifest variable expressivity such that the same pathology can present as distinct conditions in different individuals. This is known to occur in Mendelian traits but there is not a concrete basis for this phenomenon in complex traits.

It is likely that a combination of main effects, small effect variants, and interaction effects determines disease pathology. In this study we evaluated interaction effects and marginal effects of the same loci and determined that associations with disease were driven by interaction effects only. Further work to model these effects in tandem is needed to provide a more complete understanding of disease risks and mechanisms. Our findings highlight the challenges that remain in leveraging genomics in human health. A stronger emphasis on evaluating genetic interactions is needed to better understand clinical risk associations and inform personalized medicine solutions.

## Materials and Methods

### Selection of epistatic SNP pairs in the UKBB dataset

The UKBB contains genotype data for a total of 488,377 participants and electronic health records (EHRs) for nearly 400,000 participants. At the time of recruitment, participants provide information about their sociodemographic, lifestyle, and health-related factors. Physical measures (such as blood pressure and anthropometry) are also collected from all participants upon recruitment. UKBB genomic data are based on genome build GRCh37 (released in 2009). UKBB genotype data are imputed to the Haplotype Reference Consortium (HRC) panel (Bush et al., 2009).

To identify SNP pairs that were in LD due to epistatic selection, we calculated Ohta’s *D* statistics for genome-wide SNP pairs in the UKBB European ancestry population of 1000 Genomes phase III dataset (n = 503). As a first step, we calculated pairwise LD across all SNP pairs and selected the pairs with R^2^ > 0.3 using PLINK v2.0. Next, we determined which SNP pairs were in long-range LD by selecting pairs with LD that are located at least 250,000 base pairs (approximately 0.25cM) apart for intrachromosomal models. We considered all independent SNPs for each chromosome for interchromosomal models. We then filtered the results to include only the SNP pairs with strong LD by setting a threshold of R^2^ ? 0.7 and minor allele frequency > 0.1 for both variants. This yielded 186,119 unique SNPs under epistatic selection that made up 7,586,336 pairwise SNP-SNP models when considering long-range and LD. This includes 817,892 interchromosomal models. We then tested the pairwise models for epistatic selection by calculating Ohta’s *D* statistics using the *ohtadstats* R package (Petrowski et al., 2019), which provided D and D’ statistics (*D*^2^ IT, *D*^2^ IS, *D*^2^ ST, *D*′^2^ IS, and *D*′^2^ ST) for all models along with the ratios of d2is_mat to d2st_mat (ratio1) and dp2st_mat to dp2is_mat (ratio2). Finally, we selected the models for which both ratio1 and ratio2 were greater than 1, suggesting epistatic selection. This yielded 5,625,845 SNP-SNP models comprising 136,019 unique SNPs. We then tested the association of these models with phenotypes in the UKBB dataset.

### Testing of long-range epistatic SNP-SNP models for phenotypic associations

We tested the 5,625,845 SNP-SNP models for associations with 23 complex disease outcomes in the UKBB European ancestry population of unrelated individuals (n = 384,331). Phenotype definitions for each disease were based on the presence or absence of ICD9/ICD10 codes in EHRs and inclusion criteria outlined by PheCode. We conducted epistasis association tests using the FastEpistasis module in PLINK v1.9 to identify epistatic SNP-SNP models that were significantly associated with each phenotype. FastEpistasis is a software tool that computes tests of epistasis for a large number of SNP pairs as an efficient parallel extension to the PLINK epistasis module. Epistatic effects are tested by normal linear regression of a binary response on the marginal effects of each SNP and an interaction effect of the SNP pair, where SNPs are coded as additive effects, taking values 0, 1, or 2. The test for epistasis reduces to testing whether the interaction term is significantly different from zero.

### Mapping of SNP-SNP models to cytoband regions

For biological interpretability, we mapped all 5,625,845 SNP-SNP models to cytoband regions to produce cytoband-cytoband (cyto-cyto) models. Cytoband annotation was done using the UCSC Genome Browser build 37 SNP-to-cytoband map files. We also annotated the SNPs to genes using the software tool Biofilter (Bush et al., 2009). In addition to mapping the SNPs to the closest upstream or downstream gene, we manually determined which nearby genes were likely to engage in long-range interactions using the UCSC browser. We then tested the significance of the cyto-cyto models with Bonferroni correction to a threshold of 1.1× 10^−6^ based on the total number of unique cyto-cyto mappings (n = 44,860 unique cyto-cyto pairs). The 5,625,845 SNP-SNP models were binned into the unique cyto-cyto pairs, and the model in each bin with the smallest p-value meeting the Bonferroni threshold was selected as a significant cyto-cyto pair. A total of 15 cyto-cyto pairs based on the UKBB data were determined to be significant across 8 of 23 phenotypes.

### Testing for replication in the eMERGE dataset

The eMERGE is a consortium of 12 academic medical centers across the United States that have contributed EHR and genotype data of approximately 100,000 individuals into a central repository. The eMERGE phase III genomic data uses genome build GRCh37 (released in 2009). All eMERGE samples have been imputed to the HRC panel using the Michigan imputation server. We extracted phenotype data in the form of diagnosis codes from the EHRs, with samples defined as cases or controls based on the occurrence or absence of ICD9/10 code(s) grouped as per PheCode criteria. We extracted disease status from the eMERGE data for the eight phenotypes that had significant associations with epistatic SNP pairs in the UKBB data. We tested all SNP-SNP models that mapped to the 15 statistically significant cyto-cyto models for associations with 23 disease phenotypes in the eMERGE European ancestry population of unrelated individuals’ dataset. We tested all SNP-SNP models that mapped to each significant cyto-cyto model because the causal SNPs are not known. We used a Bonferroni correction based on the 15 tested cyto-cyto models (0.05 / 15). One of the 15 cyto-cyto models reached statistical significance after adjustment for multiple hypothesis testing correction with a p-value threshold of 3.5× 10^−3^.

### Main effects analysis of significant results

We tested each SNP in pairs showing significance across the eMERGE and UKBB datasets in univariate tests for association with their corresponding phenotypes. For this, we first identified proxy SNPs that were in LD (R2 > 0.5) with, and located within 1MB in either direction of, each significant SNP, using the 1000 Genomes LD panel as a reference. We then used PLINK v1.90Beta4.5 to perform a logistic regression for each SNP including sex, age, and the first five principal components as covariates. Principle components for both datasets were generated from the individuals in each dataset. A Bonferroni threshold was calculated based on the number of SNPs (the sum of the significant SNPs and the proxy SNPs) tested per phenotype. We identified potential nearby association signals using LocusZoom plots (Boughton et al., 2021).

## Data Availability

All data produced are available online. eMERGE network phase III data can be accessed through dbGaP (study ID phs001584.v2.p2). UKBB data was accessed through protocol number 32133.

## Supplementary figures

**Table S1.** Demographics of the UKBB and eMERGE case/control cohorts

|  | UKBB EUR cohort (n = 384,331) |  |  |  | eMERGE EUR cohort (n = 50,646) |  |  |  |
| --- | --- | --- | --- | --- | --- | --- | --- | --- |
|  | Cases |  | Controls |  | Cases |  | Controls |  |
|  | Female | Male | Female | Male | Female | Male | Female | Male |
| Acute Pulmonary Heart Disease | 13620 | 26750 | 197294 | 143071 | N/A | N/A | N/A | N/A |
| AD | 2301 | 2035 | 197568 | 157969 | 5096 | 4477 | 20467 | 20605 |
| COPD | 25269 | 20039 | 183855 | 147005 | N/A | N/A | N/A | N/A |
| T2D | 9923 | 15406 | 200731 | 155087 | 12871 | 12952 | 12692 | 12130 |
| Essential Hypertension | 43122 | 49825 | 164113 | 112197 | N/A | N/A | N/A | N/A |
| Fibromyalgia | 643 | 91 | 211014 | 171142 | N/A | N/A | N/A | N/A |
| Glaucoma | 4001 | 3848 | 184534 | 149911 | N/A | N/A | N/A | N/A |
| Hepatic Infection | 461 | 614 | 197049 | 158063 | N/A | N/A | N/A | N/A |
| Herpes | 194 | 158 | 211005 | 170197 | 5977 | 3271 | 19586 | 21811 |
| Hyperplasia of Prostate | 37 | 21071 | 212186 | 150283 | 119 | 12454 | 25444 | 12628 |
| Idiopathic Proctocolitis | 37095 | 28389 | 175135 | 143070 | N/A | N/A | N/A | N/A |
| Iron Deficiency Anemia | 7281 | 4324 | 205303 | 167422 | 9258 | 5465 | 16305 | 19617 |
| Ischemic Heart Disease | 13620 | 26750 | 164113 | 112197 | N/A | N/A | N/A | N/A |
| MDD | 9294 | 5695 | 190029 | 153374 | N/A | N/A | N/A | N/A |
| Cancer of Digestive Organs | 66953 | 57618 | 145479 | 114094 | N/A | N/A | N/A | N/A |
| MS | 1876 | 1283 | 194534 | 166734 | 834 | 309 | 24729 | 24773 |
| Pancreatitis | 14587 | 7029 | 189748 | 156083 | N/A | N/A | N/A | N/A |
| Psoriasis | 1771 | 1725 | 202679 | 162999 | 2485 | 1995 | 23078 | 23087 |
| SCZ | 590 | 735 | 194937 | 157428 | 638 | 475 | 24925 | 24607 |
| Tonsillitis | 104 | 81 | 212480 | 171665 | N/A | N/A | N/A | N/A |
| Ulcerative Colitis | 36834 | 27910 | 175746 | 143829 | N/A | N/A | N/A | N/A |
| Viral Infection | 296 | 397 | 210911 | 169969 | N/A | N/A | N/A | N/A |
| Breast Cancer | 14528 | 0 | 191354 | 0 | N/A | N/A | N/A | N/A |

**Fig. S1:**
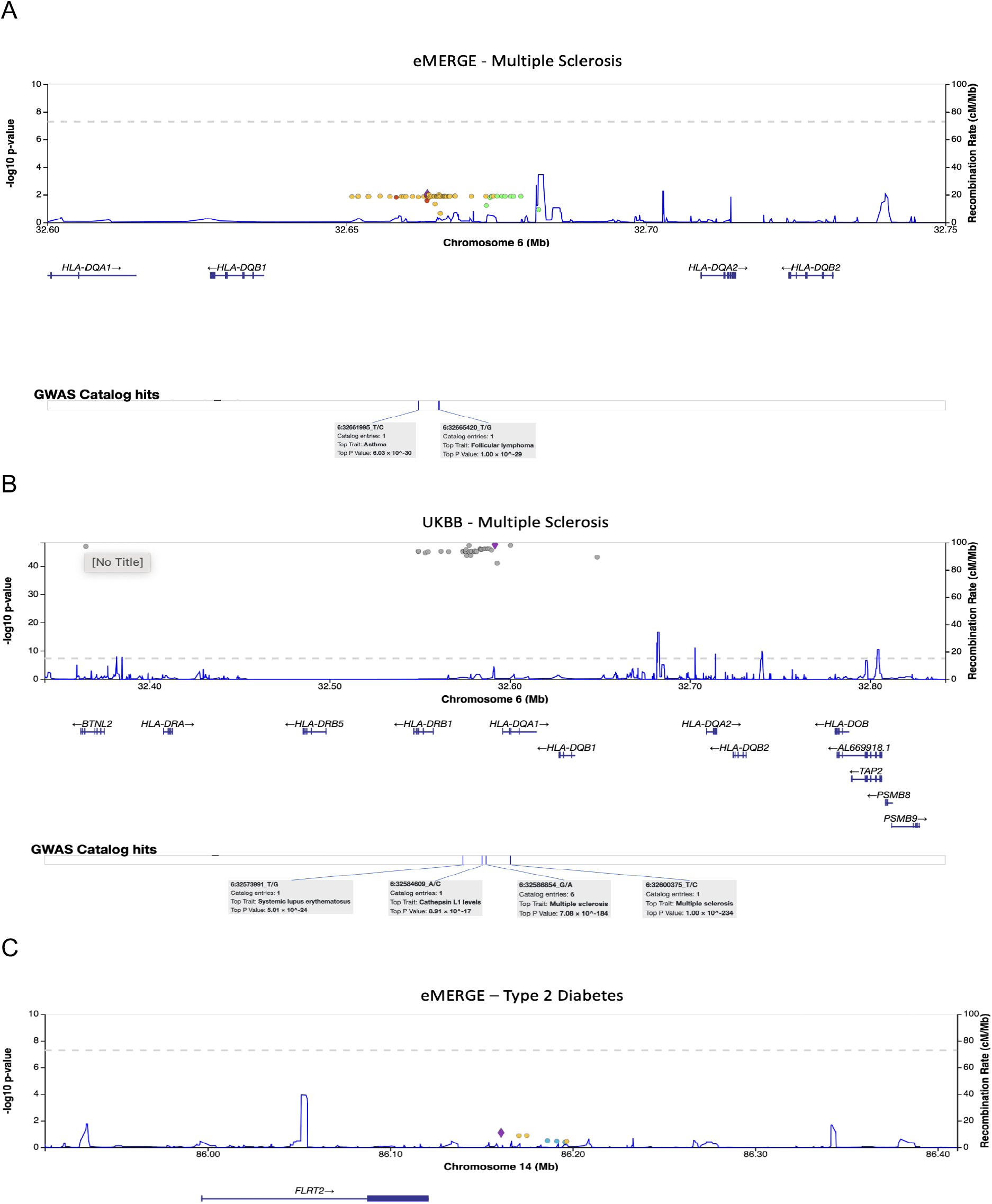
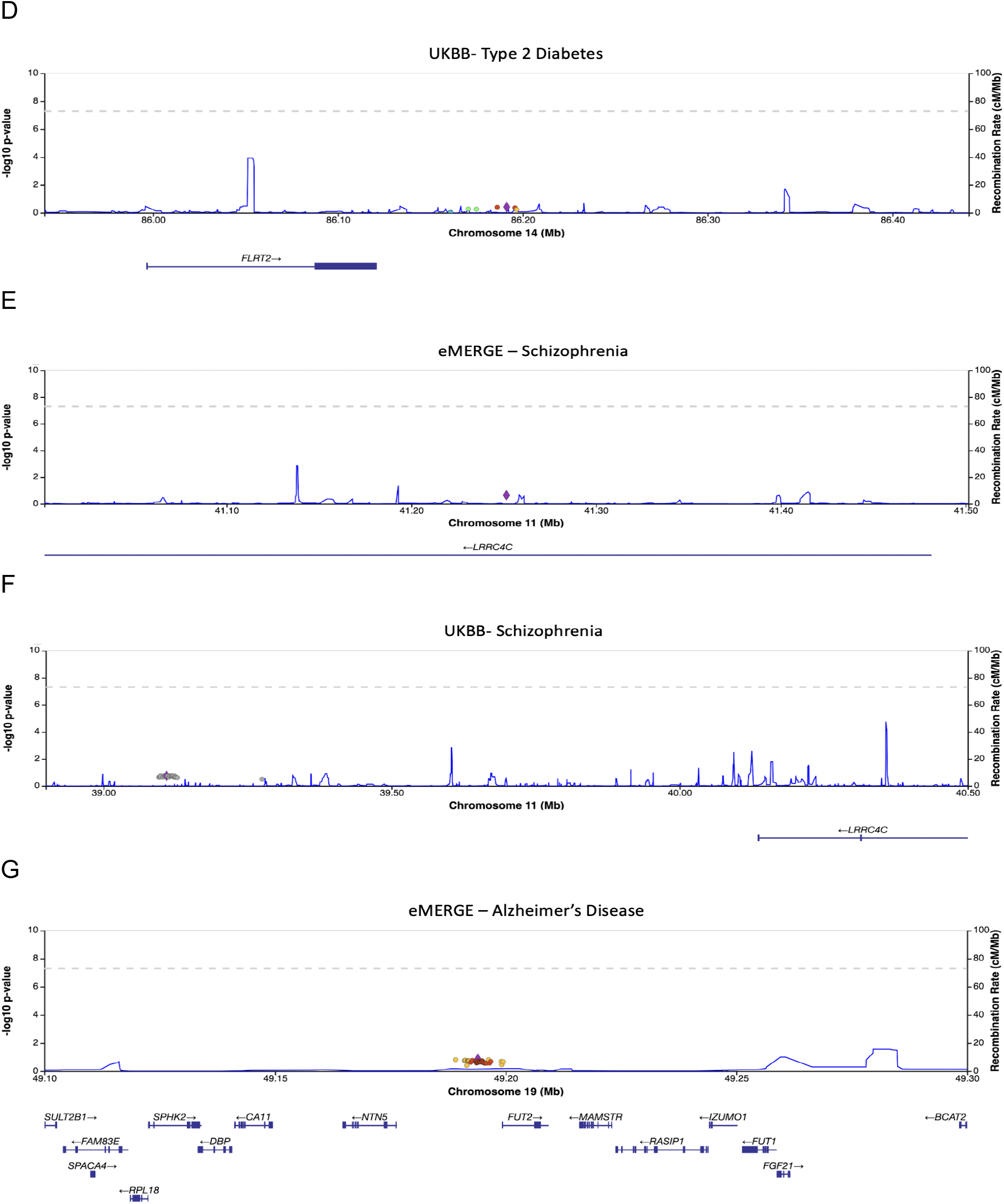
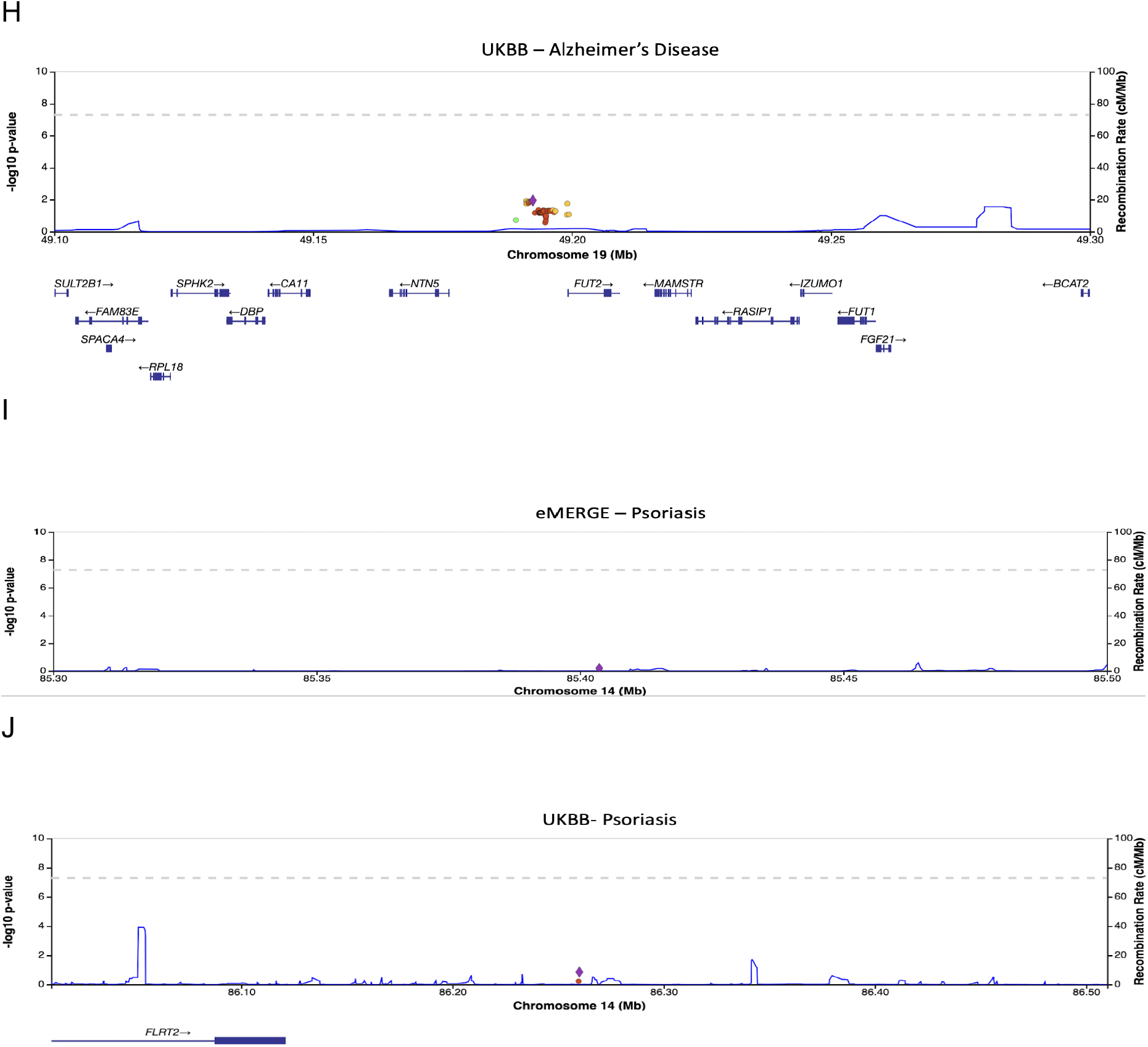
Main effect univariate analysis of significant and near-significant replicating SNPs and all their proxy SNPs (R^2^ > 0.5) within 1MB upstream or downstream

## Notes

### Competing Interest Statement

The authors have declared no competing interest.

### Funding Statement

MDR is supported by R01 HG010067 and U01 AG066833. JHM is supported by R01 LM010098 and U01 AG066833. PS is supported by F31 AG069441-01. eMERGE Network (Phase III): This phase of the eMERGE Network was initiated and funded by the NHGRI through the following grants: U01HG8657 (Group Health Cooperative/University of Washington); U01HG8685 (Brigham and Womens Hospital); U01HG8672 (Vanderbilt University Medical Center); U01HG8666 (Cincinnati Childrens Hospital Medical Center); U01HG6379 (Mayo Clinic); U01HG8679 (Geisinger Clinic); U01HG8680 (Columbia University Health Sciences); U01HG8684 (Childrens Hospital of Philadelphia); U01HG8673 (Northwestern University); U01HG8701 (Vanderbilt University Medical Center serving as the Coordinating Center); U01HG8676 (Partners Healthcare/Broad Institute); and U01HG8664 (Baylor College of Medicine).

### Author Declarations

Institutional Review Board of the University of Pennsylvania gave ethical approval for this work through IRB Protocol #850838. The project qualified as exempt.

